# Prevalence and Factors Associated With Late Antenatal Care Booking Among Pregnant Women at Magoye Rural Health Center in Mazabuka District, Southern Province, Zambia: A Facility-Based Cross-Sectional Study

**DOI:** 10.1101/2025.08.08.25333288

**Authors:** Mervis Nakacha Mweene Chilanga

**Author notes:** Corresponding author: Mervis Nakacha Mweene Chilanga.

## Abstract

**Background:** Late antenatal care (ANC) booking has continued to be a public health issue among women of childbearing age, 15 to 49 years. It is known to limit the number of ANC contacts and compromise the quality of ANC services due to delayed screening and diagnosis for pregnancy complications. The Ministry of Health and its partners have implemented interventions to scale up ANC services; however, many women still book later than recommended. Evidence of late ANC from rural primary health facilities of the Southern Province is scarce. This study estimated the prevalence of late ANC booking and its associated factors among pregnant women at Magoye Rural Health Center.

**Methodology:** A facility-based cross-sectional study was conducted from 9 June to 19 August 2024 among pregnant women attending antenatal care at Magoye Rural Health Center, Mazabuka District, Southern Province, Zambia. Women aged 15 to 49 years were recruited consecutively and interviewed using a structured questionnaire (n=355, response rate=95.9%). Late ANC booking was defined as first ANC booking after 14 weeks of gestation in line with the Zambian Ministry of Health policy. Bivariable and multivariable logistic regression were used to determine factors associated with late ANC booking. Variables with P < 0.25 in the bivariable analysis were entered into the multivariable model. The results are reported as adjusted odds ratios (AOR) with 95% confidence intervals (CI).

**Results:** The prevalence of late ANC booking was 68.5% (243/355; 95% CI: 63.4-73.1). In the multivariable model, the odds of late booking were higher among married than unmarried women (AOR: 3.96, 95% CI: 2.24-7.00), women who had given birth before than those who had not (AOR: 3.11, 95% CI: 1.61-6.00), and women not in government employment than government employees (AOR: 4.34, 95% CI: 1.28-14.75). Women older than 19 years also had higher odds than younger women (AOR: 2.01, 95% CI: 1.01-4.01).

**Conclusion:** The prevalence of late ANC booking was found to be high. Late ANC booking was associated with being married, having had previous births, not being in government employment and being older than 19 years. Since the sample was non-random and drawn from one facility, the prevalence applies to women attending this facility during the study period and not to all pregnant women in the district. There is a need to test strategies that encourage timely ANC booking among pregnant women with previous births and those that involve male partners.

## Introduction

Antenatal care (ANC) is defined as a systematic assessment and anticipatory guidance of a pregnant woman (MoH, 2022). Pregnancy is one of the most important periods in the life of a woman, a family, and society (World Health Organization [WHO], 2016). During pregnancy, women undergo many physiological changes that can lead to life-threatening problems for both the expectant mother and the baby, leading to a compromise in maternal health. Antenatal care is one of the key indicators of maternal health, as most maternal and neonatal mortalities and morbidities can be prevented through ANC. Antenatal care allows for improvement in maternal health and the survival of the baby, as well as providing information on warning signs, nutrition, contraception, HIV/AIDS, and infant feeding (Pattinson and Buchmann, 2017). During ANC, screening of common pregnancy-related ailments like preeclampsia, fetal abnormalities, hypertension, and other pregnancy-related conditions is done.

A pooled analysis of Demographic and Health Surveys from 16 sub-Saharan African countries (2018-2022) found that 53.8% (95% CI 46.2 to 61.3) of women started ANC after the first trimester, ranging from 27.8% in Liberia to 75.8% in Nigeria (Mare et al., 2025).

Antenatal care-seeking behaviors among pregnant women have remained poor, as it is usually started late, despite it being free (Mare et al., 2025). WHO recommends that the first ANC contact take place within the first 12 weeks of gestation (WHO, 2016). Zambian national guidelines define early booking as a first visit at or before 14 weeks, and late booking as a visit after 14 weeks (MoH, 2012; Sinyange et al., 2016). This study used the national 14-week threshold. Estimates using a 12-week threshold are therefore not directly comparable. Zambia has aligned itself with the WHO 2030 target on maternal health of reducing the maternal mortality ratio to less than 70 deaths per 100,000. In the quest to resolve the issues surrounding maternal health concerning antenatal care services, the Ministry of Health has put in place policies, among which are the following: eight (8) ANC contacts during pregnancy, early ANC booking; 14 weeks or earlier, provision of ANC services by skilled health personnel and free ANC services (MoH, 2022).

It is recommended that pregnant women have 8 ANC contacts according to the ANC model by the World Health Organization (WHO, 2016). During the subsequent contacts, pregnant women should have an ultrasound done by the 24^th^ week of gestation to ascertain the true age of the pregnancy (WHO, 2016). The WHO ANC model shows a shift from the old focused ANC model (FANC model), which only recommended 4 visits during one’s gestation period.

Other initiatives for scaling up ANC services include: health education, nutrition counseling, HIV testing and counseling, malaria prevention, blood pressure testing, and urine testing all to be done during ANC visits. Most evidence on late ANC booking in Zambia comes from national surveys or urban and peri-urban sites. Little is known about booking timing at rural primary health facilities in Southern Province, where facility-level evidence is needed to target local interventions. This study estimated the prevalence of late ANC booking, using the national 14-week definition, and identified its associated factors among women attending Magoye Rural Health Center in Mazabuka District in 2024.

While determinants of late ANC booking such as multiparity and marital status are documented globally, empirical evidence specific to rural primary health facilities in Southern Province, Zambia, remains scarce. Sub-district implementation of maternal health policies requires context-specific evidence to identify and address local structural and sociocultural barriers that national surveys like the Zambia Demographic and Health Survey (ZDHS) may not adequately capture. Conducting facility-level research in a high catchment rural feeder facility provides necessary baseline empirical data to tailor local health policy and intervention strategies.

The objective of this study was to determine the prevalence and factors associated with late antenatal care booking among pregnant women at Magoye Rural Health Center in Mazabuka District, Southern Province, Zambia.

## Methods

### Study design, setting, and population

We conducted a facility-based analytical cross-sectional study. The design cannot establish temporal order, so associations are not interpreted as causal.

Magoye Rural Health Center was chosen because it is a rural primary-level facility that feeds Mazabuka General Hospital, serves a catchment population of 8,740, and was accessible during the study period. The site was therefore selected for feasibility, and the findings are interpreted as applying to this facility.

### Study population

The study population was all pregnant women aged 15 to 49 years who attended ANC at Magoye Rural Health Center during the study period.

### Inclusion and exclusion criteria

Women were included if they were aged 15 to 49 years, lived in the facility catchment area and consented. Women too ill to be interviewed were excluded.

### Sample Size Determination

The sample size was calculated with Cochran’s formula for a single proportion, n = Z²p (1 − p)/d², with Z = 1.96, d = 0.05, and an expected prevalence of 0.70. This gives 323 women. Adding 10% for non-response gave 355. This is close to the 72.0% reported in the Copperbelt Province (Banda, 2012) and lower than the 86.6% reported in Ndola (Chewe, 2015). The sample size was calculated for estimating prevalence and not for the regression analysis.

### Participants and sampling

All pregnant women aged 15 to 49 years who attended the ANC clinic during the study period (9 June to 19 August 2024), lived in the facility catchment area and gave consent (or assent with parental consent for ages 15 to 17) were approached consecutively on each clinic day. Of 370 eligible women approached, 15 declined (4.1%) due to lack of time to answer the questionnaire, giving a response rate of 95.9% and a final sample of 355.

### Data collection

Structured questionnaires (English and Tonga) were used to collect data from pregnant women. Four trained research assistants helped collect data to reduce interviewer bias.

### Study variables

#### Outcome variable

Gestational age at the first ANC contact was calculated from the last menstrual period reported by the woman, or from clinical palpation when the date was unknown, and was recorded in completed weeks. A woman who first attended at 14 weeks and 0 to 6 days was therefore recorded as 14 weeks and classed as an early booker. Late ANC booking was defined as a first ANC contact after 14 completed weeks of gestation in line with the Ministry of Health definition (MoH, 2012), and was coded 0 (early, ≤14 weeks) or 1 (late, >14 weeks).

#### Independent variables

Socio-demographic variables were age in completed years (≤19 years, >19 years), marital status (single, married, divorced, widowed), highest level of education attained (none, primary, secondary, tertiary), occupation (government employed, private employed, self-employed, not employed), monthly income in Zambian kwacha (K500 or less, K501–K1000, above K1000), whether the pregnancy was planned (yes, no) and usual mode of transport to the clinic (foot, bicycle, bus, other). Women aged 15 to 19 years were classed as adolescents, in line with the World Health Organization definition of adolescence as 10 to 19 years (WHO, n.d.).

Obstetric variables were gravidity (primigravida, multigravida), parity (nulliparous, parous), and self-reported history of miscarriage, stillbirth, caesarean birth and other complications in previous pregnancies (each yes or no).

For the regression analysis, categories with few women were combined: marital status as married versus not married (single, divorced and widowed); education as primary or none versus secondary or higher; occupation as government employed versus other; and income as ≤K1000 versus >K1000. Age, whether the pregnancy was planned, mode of transport, parity and history of miscarriage were analyzed in the categories listed above. Tables 1 and 2 show the original categories, and Tables 3 and 4 show the groupings used in the regression models.

**Table 1.**
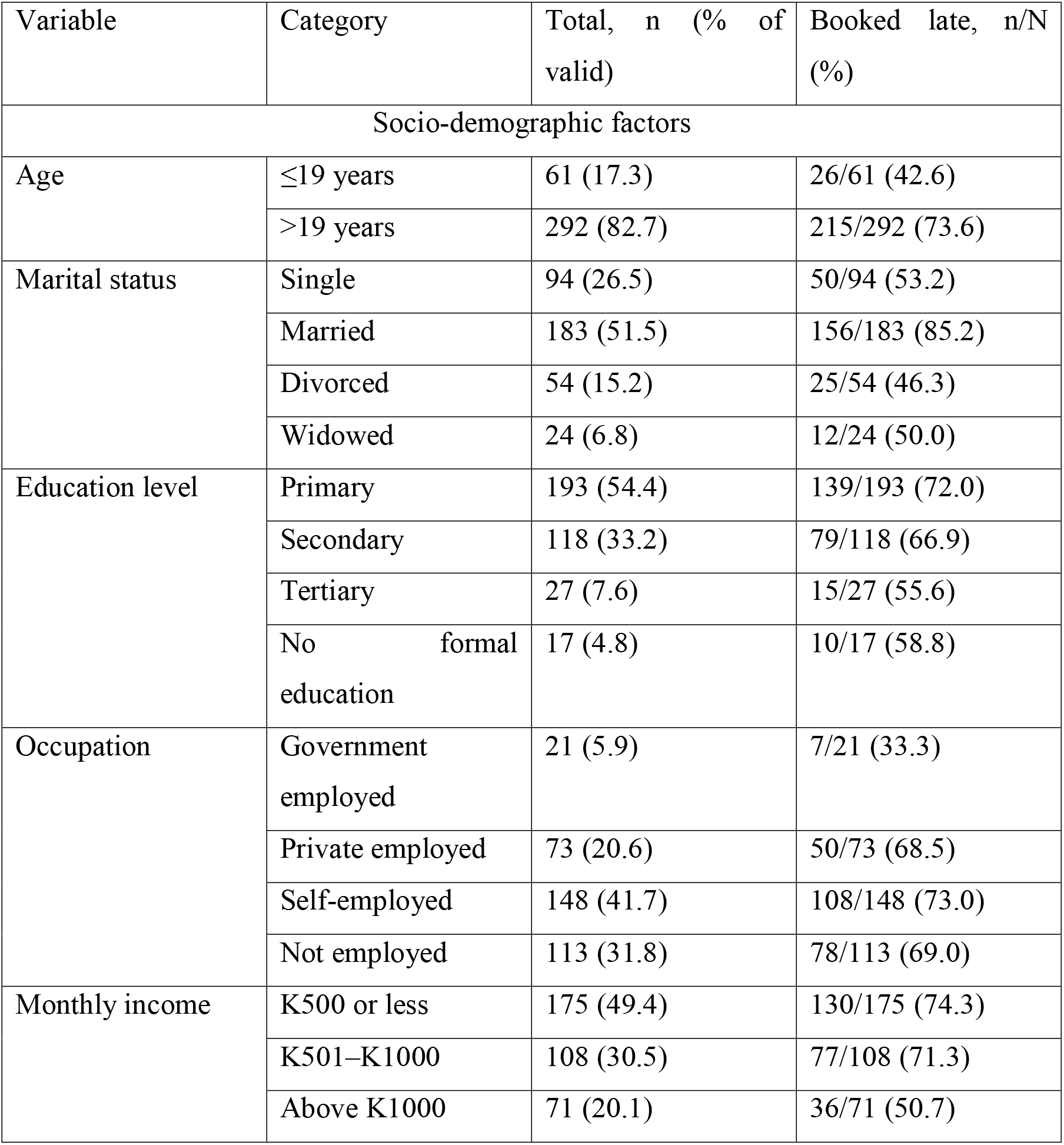
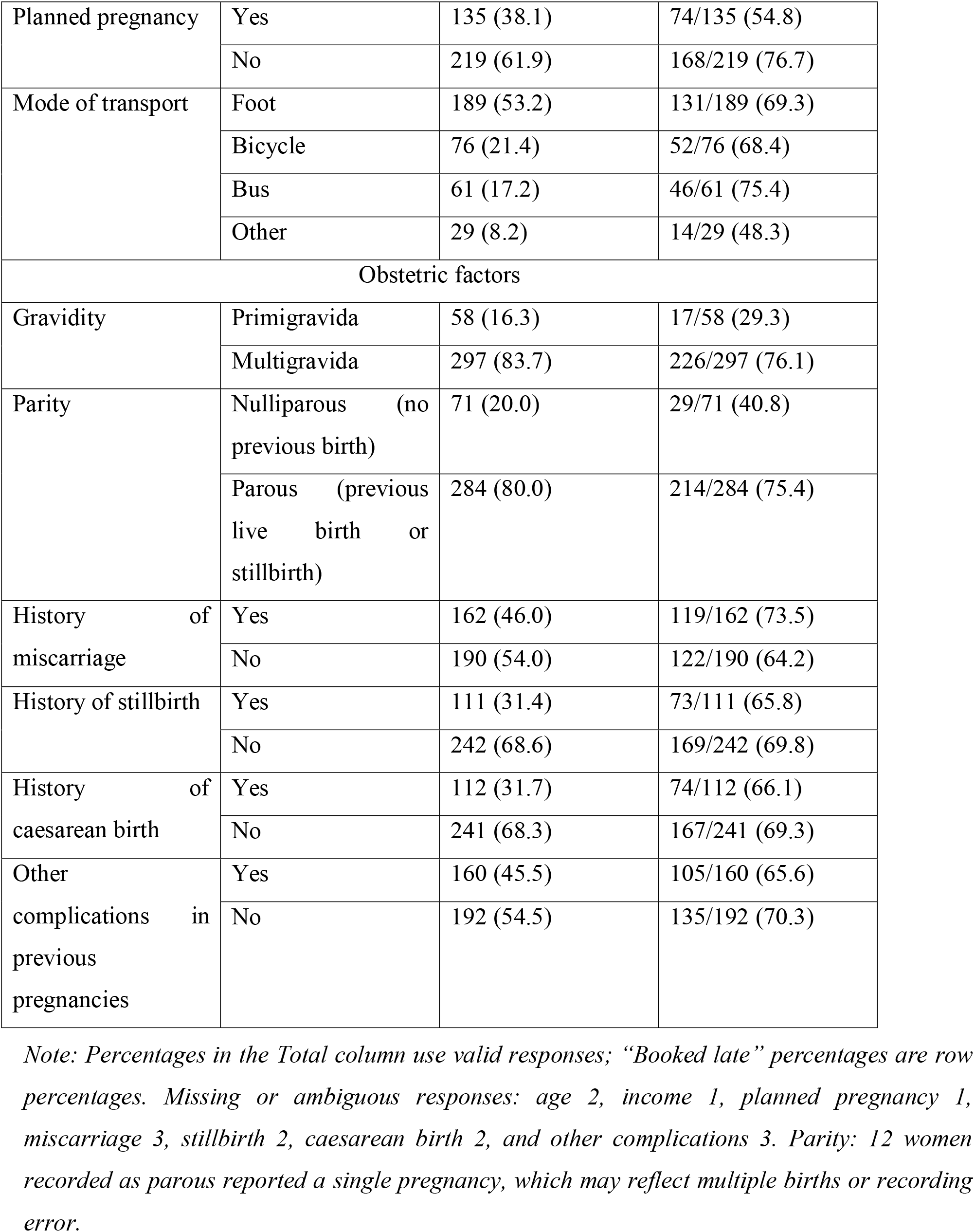
Participant characteristics and proportion booking late (N = 355)

**Table 2.**
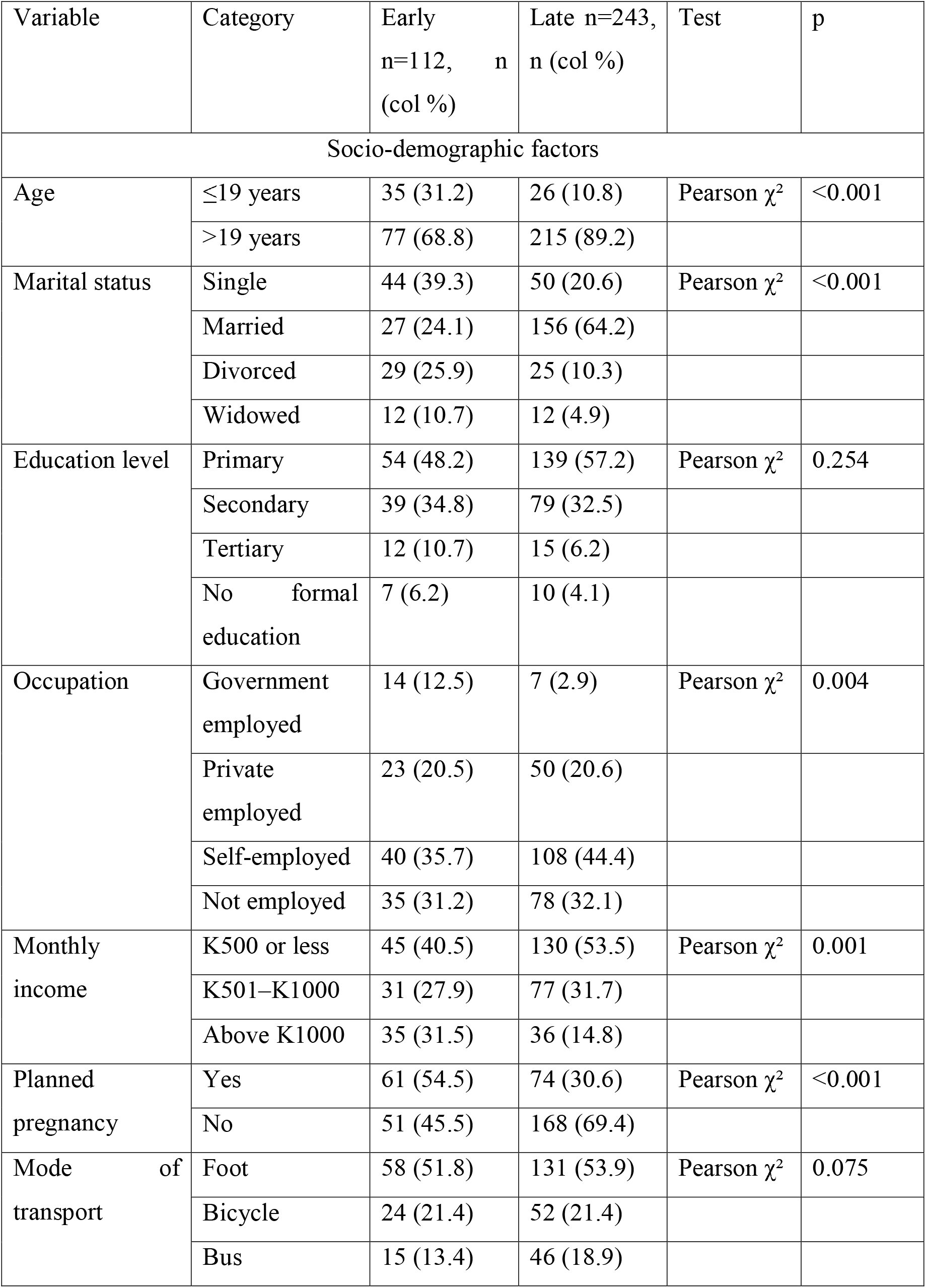
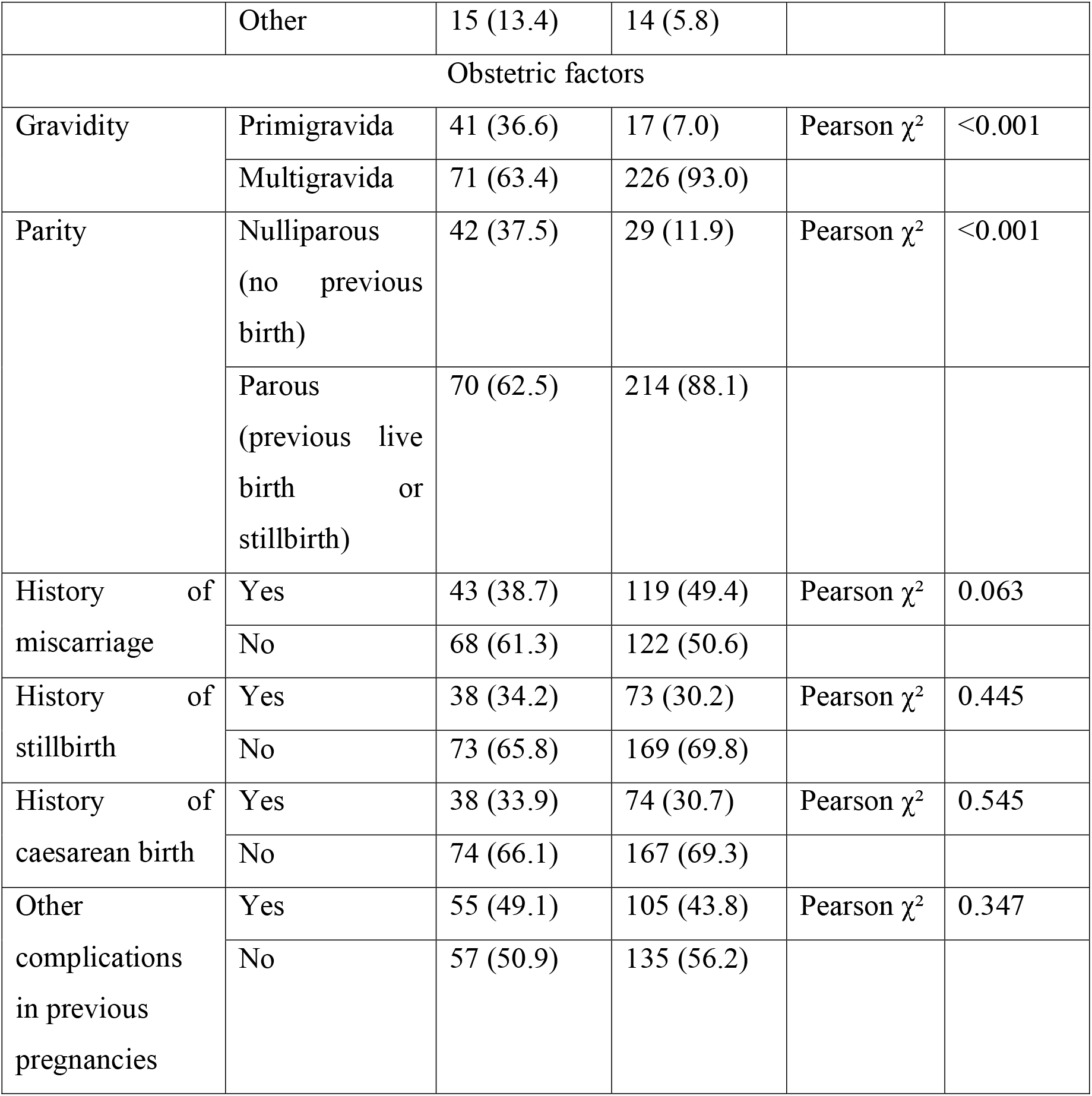
Characteristics of early (≤14 weeks) and late (>14 weeks) bookers.

**Table 3:** Bivariable (crude) logistic regression.

| Variable | Category | COR (95% CI) | p |
|---|---|---|---|
| Age | ≤19 years (ref) | 1.00 |  |
|  | >19 | 3.76 (2.12–6.65) | <0.001 |
| Marital status | Not married (ref) | 1.00 |  |
|  | Married | 5.64 (3.40–9.37) | <0.001 |
| Education | Primary or none (ref) | 1.00 |  |
|  | Secondary or higher | 0.75 (0.48–1.19) | 0.223 |
| Occupation | Government employed (ref) | 1.00 |  |
|  | Other | 4.82 (1.89–12.30) | 0.001 |
| Income | ≤K1000 (ref) | 1.00 |  |
|  | >K1000 | 0.38 (0.22–0.64) | <0.001 |
| Planned pregnancy | Yes (ref) | 1.00 |  |
|  | No | 2.72 (1.71–4.31) | <0.001 |
| Transport | Foot (ref) | 1.00 |  |
|  | Bicycle | 0.96 (0.54–1.70) | 0.887 |
|  | Bus | 1.36 (0.70–2.63) | 0.364 |
|  | Other | 0.41 (0.19–0.91) | 0.029 |
| Gravidity | Primigravida (ref) | 1.00 |  |
|  | Multigravida | 7.68 (4.11–14.34) | <0.001 |
| Parity | Nulliparous (ref) | 1.00 |  |
|  | Parous | 4.43 (2.57–7.63) | <0.001 |
| Miscarriage | No (ref) | 1.00 |  |
|  | Yes | 1.54 (0.98–2.44) | 0.064 |
| Stillbirth | No (ref) | 1.00 |  |
|  | Yes | 0.83 (0.51–1.34) | 0.445 |
| Caesarean | No (ref) | 1.00 |  |
|  | Yes | 0.86 (0.54–1.39) | 0.545 |
| Other complications | No (ref) | 1.00 |  |
|  | Yes | 0.81 (0.51–1.26) | 0.347 |

**Table 4.** Multivariable logistic regression (n = 348; 238 late, 110 early)

| Variable | Category | AOR (95% CI) | p |
|---|---|---|---|
| Age | ≤19 years (ref) | 1.00 |  |
|  | >19 | 2.01 (1.01–4.01) | 0.047 |
| Marital status | Not married (ref) | 1.00 |  |
|  | Married | 3.96 (2.24–7.00) | <0.001 |
| Education | Primary or none (ref) | 1.00 |  |
|  | Secondary or higher | 0.94 (0.53–1.66) | 0.825 |
| Occupation | Government employed (ref) | 1.00 |  |
|  | Other | 4.34 (1.28–14.75) | 0.019 |
| Income | ≤K1000 (ref) | 1.00 |  |
|  | >K1000 | 0.69 (0.33–1.47) | 0.342 |
| Planned pregnancy | Yes (ref) | 1.00 |  |
|  | No | 1.72 (1.00–2.97) | 0.049 |
| Transport | Foot (ref) | 1.00 |  |
|  | Bicycle | 0.99 (0.50–1.94) | 0.966 |
|  | Bus | 1.32 (0.61–2.86) | 0.487 |
|  | Other | 0.80 (0.31–2.08) | 0.647 |
| Parity | Nulliparous (ref) | 1.00 |  |
|  | Parous | 3.11 (1.61–6.00) | <0.001 |
| Miscarriage | No (ref) | 1.00 |  |
|  | Yes | 0.84 (0.47–1.50) | 0.552 |

*Hosmer–Lemeshow χ² = 13.32, df = 8, p = 0.101; maximum VIF 1.56; 10 early bookers per parameter. Variables entered: those with p < 0.25 in Table 3 (age, marital status, education, occupation, income, planned pregnancy, transport, parity, miscarriage). Gravidity was not entered because it overlaps with parity (272 of 284 parous women were multigravida).*

#### Operational definitions

ANC booking: the first ANC contact for the current pregnancy.

Late ANC booking: a first ANC contact after 14 completed weeks of gestation.

Gravidity: the number of pregnancies a woman has had, including the current one. Women were classed as primigravida (first pregnancy) or multigravida (more than one pregnancy).

Parity: having given birth previously (live birth or stillbirth), classified as nulliparous (none) or parous (one or more).

Planned pregnancy: a pregnancy the woman reported she wanted at the time she became pregnant.

History of miscarriage, stillbirth, caesarean birth and other complications in previous pregnancies: self-reported by the woman and recorded as yes or no.

#### Data management

Data were collected on paper questionnaires and entered into Microsoft Excel for coding, with data validation rules used to reduce entry errors. After cleaning and validation, the coded dataset was exported to IBM SPSS Statistics version 29 for analysis. The dataset was password-protected to maintain confidentiality, and a secure backup copy was stored on Google Drive to guard against loss from device failure.

#### Missing data

Missing or ambiguous responses were few: age (2), monthly income (1), planned pregnancy (1), history of miscarriage (3), history of stillbirth (2), history of caesarean birth (2) and other complications in previous pregnancies (3) (Table 1 note). A woman with a missing or ambiguous value was excluded from analyses involving that variable (complete-case analysis). The multivariable model therefore used the 348 women with complete data on all entered variables.

### Statistical Analysis

Data were analyzed with IBM SPSS Statistics version 29. Categorical variables were summarized as frequencies and percentages, and the prevalence of late booking was reported with its 95% confidence interval. The 95% confidence interval for the prevalence was calculated using the Wilson score method. Associations between each characteristic and late booking were first examined using Pearson’s chi-square test. Crude odds ratios (COR) were estimated by bivariable logistic regression. Variables with p < 0.25 in bivariable analysis were entered into the multivariable model. Gravidity met this criterion but was not entered because it orerlaps with parity (272 of 284 parous women were multigravida); only parity was retained. Some categories were combined for the regression analysis, as described under study variables. Results are presented as adjusted odds ratios (AOR) with 95% confidence intervals, and p < 0.05 was taken as statistically significant.

Multicollinearity was assessed with variance inflation factors (VIF), and a value above 5 was taken to indicate a problem. Model fit was assessed with the Hosmer-Lemeshow test, and a p-value above 0.05 was taken to show no evidence of poor fit.

### Ethical considerations

The study was conducted in accordance with the principles of the Declaration of Helsinki. It was approved by the University of Zambia Biomedical Research Ethics Committee (UNZABREC; REF. No. 5212-2024) and the National Health Research Authority. Permission to conduct the study was obtained from the Mazabuka District Health Office. Women aged 18 years and above gave written informed consent. For girls aged 15 to 17 years, written parental or guardian consent was obtained first, and the girl then gave written assent. In both cases, participants who could not write gave verbal consent with a thumbprint. Assent was explained in simple language and in the local language by a trained research assistant, who asked each girl to say in her own words what the study involved before she signed. Interviews were conducted in an office space within the facility. Questionnaires carried a unique code to ensure anonymity of respondents. Paper questionnaires were kept in a locked mini-mobile trunk. Electronic data were password-protected and accessible only to the researcher. Participation was voluntary, and women could withdraw at any time without affecting the care they received.

### Dissemination of the Research Findings

Reports of the research findings were shared with the Provincial Health Office (PHO), the District Health Office (DHO), the National Health Research Authority (NHRA), and the School of Public Health at the University of Zambia.

## Results

### Participant characteristics

A total of 355 women were enrolled in the study, most of whom were older than 19 years (82.7%), married (51.5%), had primary education only (54.4%) and were self-employed (41.7%). Nearly half (49.4%) earned K500 or less a month, and most had unplanned pregnancies (61.9%), walked to the clinic (53.2%), were multigravida (83.7%), and were parous (80.0%). Table 1 gives the full distribution.

### Prevalence of late ANC booking

Of the 355 women that entered the study, 243 (68.5%; 95% CI 63.4 to 73.1) booked after 14 weeks of gestation and 112 (31.5%) booked at or before 14 weeks.

Table 2 compares the 112 early bookers with the 243 late bookers. Percentages are column percentages, meaning the share of each group with that characteristic. Late bookers were more often older than 19 years (89.2% vs 68.8%) and married (64.2% vs 24.1%), while single women made up 20.6% of late bookers and 39.3% of early bookers. Government-employed women were 2.9% of late bookers and 12.5% of early bookers. An unplanned pregnancy was reported by 69.4% of late bookers and 45.5% of early bookers. Women earning more than K1000 a month made up 14.8% of late and 31.5% of early bookers. Primary education only was more common among late bookers (57.2% vs 48.2%), and tertiary education was less common (6.2% vs 10.7%), but the overall association with education was not significant (p = 0.254). Multigravida women were 93.0% of late bookers and 63.4% of early bookers, and women who had given birth before (parous) were 88.1% and 62.5%, respectively. Mode of transport did not differ significantly (p = 0.075): 53.9% of late and 51.8% of early bookers walked to the clinic. In bivariable analysis, late booking was associated with older age (COR: 3.76, 95% CI: 2.12-6.65), being married (COR: 5.64, 95% CI: 3.40-9.37), non-government employment (COR: 4.82, 95% CI: 1.89-12.30), an unplanned pregnancy (COR 2.72; 95% CI: 1.71-4.31), multigravidity (COR: 7.68, 95% CI: 4.11-14.34) and parity (COR: 4.43, 95% CI: 2.57-7.63). Income above K1000 was associated with lower odds of late booking (COR: 0.38, 95% CI: 0.22-0.64).

In the multivariable model, four factors remained independently associated with late booking. Married women had higher odds than unmarried women (AOR: 3.96, 95% CI: 2.24-7.00). Women who had given birth before had higher odds than women who had not (AOR: 3.11, 95% CI: 1.61-6.00). Women not in government employment had higher odds than government employees (AOR: 4.34, 95% CI: 1.28-14.75), although this estimate is imprecise because only 21 women were government-employed. Women older than 19 years had higher odds than younger women (AOR: 2.01, 95% CI: 1.01-4.01), and the lower confidence limit is close to 1. An unplanned pregnancy was borderline (AOR: 1.72, 95% CI: 1.00-2.97). Education, income, mode of transport, and history of miscarriage were not significantly associated with late booking after adjustment (Table 4). Gravidity was not entered into the model because it overlaps with parity (272 of the 284 parous women were multigravida). Because late booking was common (68.5%), odds ratios overstate the corresponding relative risks and should be read as showing the direction and rough strength of association. The model showed no evidence of poor fit (Hosmer- Lemeshow χ² = 13.32, df = 8, p = 0.101), and all VIF values were below 1.6.

## Discussion

The prevalence of late ANC booking in the study was 68.5% among women attending Magoye Rural Health Center. Married women, women who had given birth before, women not in government employment, and women older than 19 years had higher odds of late booking, and an unplanned pregnancy was of borderline significance. The finding sits against national survey data showing that nearly all Zambian women attend ANC at least once (Zambia Statistics Agency, Ministry of Health and ICF, 2019), yet a smaller share start in the first trimester. At this facility, the problem is therefore the timing of the first contact and not whether women reach the clinic.

The prevalence of 68.5% is lower than the 72.0% reported in rural and urban communities of Copperbelt Province (Banda, 2012), the 86.6% reported in Ndola (Chewe, 2015) and the 85.3% in Kigamboni, Dar es Salaam (Ndomba et al., 2023). The rural setting of this study is consistent with South African evidence that late presentation is more common in rural than in peri-urban communities (Ebonwu et al., 2018). Differences between studies may reflect differences in the definition of late booking (14 weeks in this study, whereas Ndomba et al. (2023) classed booking at week 13 or later as late, a stricter cutoff that partly explains their higher figure), in setting, and in how gestational age was estimated, so comparisons between studies need care. Of these studies, Banda (2012) and Ebonwu et al. (2018) included rural communities making them the closest comparisons with the present rural facility.

Married women had higher odds of late booking than unmarried women (AOR: 3.96, 95% CI: 2.24-7.00), consistent with Chewe (2015) in Ndola and with Ebonwu et al. (2018), who found about twice the odds among married women in a rural South African community. Marital status was not reported as a significant factor in the Zambia Demographic and Health Survey analysis (Sinyange et al., 2016). The crude proportions booking late were similar among single (53.2%), divorced (46.3%) and widowed (50.0%) women and far higher among married women (85.2%) (Table 1). In the regression analysis, the first three groups were combined as “not married”. Qualitative studies elsewhere point to partner influence on the timing of ANC (Olufemi Olayinka et al., 2022; Warri and George, 2020). However, partner involvement in decision-making and who controls transport money were not measured in this study, so it cannot be ascertained why married women booked later. This is a hypothesis that needs testing in this setting.

Women who had given birth before had about three times the odds of late booking compared with women who had not (AOR: 3.11, 95% CI: 1.61-6.00). This agrees with Towongo et al. (2022) in Uganda, where women with four or more children were less likely to book in the first trimester, and with Dorji et al. (2019) in Bhutan, where late booking was more common among women with previous pregnancies. Gravidity and parity are closely related in this study, so only parity was entered in the model, and their separate effects cannot be distinguished. One possible explanation, evident from previous studies, is that women with previous pregnancies feel more experienced and see early care as less necessary. Providers in South Africa described multigravida women postponing ANC because of their experience (Jinga et al., 2019), and mothers in Bhutan expressed the same view (Dorji et al., 2019). Perceived risk and confidence were not measured in this study. Women not in government employment had higher odds of late booking than government employees (AOR: 4.34, 95% CI: 1.28-14.75). This estimate is imprecise because only 21 women were government-employed, so it should be read cautiously. The crude figures suggest that the difference relates to government employment specifically and not to employment in general: the proportion booking late was similar among private employees (68.5%), self-employed women (73.0%), and unemployed women (69.0%), and much lower among government employees (33.3%) (Table 1). Income did not explain the difference, because it was not independently associated with late booking after adjustment. One possible explanation is that government employment brings paid leave and maternity benefits that make early attendance easier. In South Africa, providers reported that employed women without maternity benefits or paid leave gave priority to work over clinic visits (Jinga et al., 2019). Studies that compared employed with unemployed women reached different conclusions; for example, higher odds of late presentation among employed women in a rural South African community (Ebonwu et al., 2018), so the findings are not directly comparable. Job conditions, access to leave and health knowledge were not measured in this study.

Women older than 19 years had about twice the odds of late booking compared with women aged 19 years or younger (AOR: 2.01, 95% CI: 1.01-4.01). The lower confidence limit is close to 1, and only 61 women were in the younger group, so this finding is weak. It is the opposite of most earlier work, where younger women booked later: adolescents had higher odds of late booking in the Zambia Demographic and Health Survey (Sinyange et al., 2016) and in rural South Africa (Ebonwu et al., 2018), and women aged 15 to 24 had higher odds across sub- Saharan Africa (Mare et al., 2025). Older women in this sample were more often parous, but age remained associated with late booking after parity was included in the model, so parity alone does not explain it. Providers in South Africa also described older women as the group that postpones booking: “The young ones, most of the time they book early, the problem is the old ones” (Jinga et al., 2019). Why older women booked later was not measured here, so this remains a hypothesis that needs testing in a larger sample that includes more adolescents.

An unplanned pregnancy was associated with higher odds of late booking, although the evidence is borderline (AOR: 1.72, 95% CI: 1.00-2.97; p = 0.049). The direction agrees with earlier studies. In South Africa, unplanned pregnancy was an independent risk factor for late presentation in rural and peri-urban communities (Ebonwu et al., 2018), and in the Zambia Demographic and Health Survey, women who wanted their last child later or wanted no more children had higher odds of booking late (Sinyange et al., 2016). Qualitative studies describe how an unplanned pregnancy can delay booking: women may keep the pregnancy secret, struggle to accept it, fear stigma or the reaction of their parents, or take time to decide whether to continue it (Jinga et al., 2019; Warri and George, 2020). These reasons were not measured here. Because most women in this study (61.9%) reported an unplanned pregnancy (Table 1), linking family planning services with messages on early booking may be worth testing.

These findings come from one facility, and they suggest where the 14-week early-booking target is most likely to be missed: among women who already have children and among married women. The MoH and its partners promote early booking, free ANC, and a minimum of eight contacts (MoH, 2022; WHO, 2016). A woman who first attends at 20 weeks has missed the first contact in the WHO schedule and can attend at most seven of the eight scheduled contacts. Facility and community programs could use postnatal, immunization and family planning contacts to reach women who have already had children, and could include partners in routine health messages. These ideas need testing and were not evaluated here.

### Study strengths and limitations

This study provides facility-level data from a rural primary health facility in Southern Province, where evidence on the timing of antenatal booking is scarce. Women were recruited consecutively over 10 weeks with a response rate of 95.9%; the questionnaire was administered in English and Tonga by trained data collectors, and missing data were few. Multicollinearity and model fit were assessed, and the findings are reported with their confidence intervals so that imprecise estimates are visible. However, the study has limitations. The cross-sectional design shows association only, so no causal inference can be drawn. Sampling was non-random and limited to women attending one rural facility during 10 weeks, so the 68.5% prevalence applies to women attending this facility in that period and not to all pregnant women in the catchment area or Mazabuka District. Women who never attend ANC were not included, and they are likely to book later or not at all, so the true district prevalence of late or no booking could be higher. Gestational age was estimated from the last menstrual period reported by the woman, which is subject to recall error, or from clinical palpation, without first-trimester ultrasound, so some women may have been misclassified. There were only 112 early bookers, so the multivariable model had about 10 early bookers per parameter, and some estimates, notably for occupation (21 government-employed women), are imprecise. Finally, partner influence, distance, cost, knowledge and perceptions of ANC were not measured, and so residual confounding is likely.

### Recommendations

The facility should record gestational age at the first ANC contact for every woman and report each quarter the proportion who book at or before 14 weeks, so that progress against the national early-booking target can be monitored at facility and district level.

Women who have already given birth, who made up 88% of the women booking late, can be reached through postnatal, child immunization and family planning contacts. Staff at these contacts should encourage women to book their next pregnancy early, and should present early booking as a service women are entitled to, without describing previous pregnancies or births as a risk.

Married women were more likely to book late. Facilities, community health workers and Safe Motherhood Action Groups could pilot the invitation of male partners to ANC-related talks and community discussions on the timing of the first contact, and should measure whether booking earlier follows.

Because the reasons behind these associations were not measured, qualitative and multi-site studies in Southern Province are needed to examine partner involvement, distance and cost, knowledge, and women’s perceptions of early booking.

### Conclusion

About two in three pregnant women attending Magoye Rural Health Center booked for ANC after 14 weeks of gestation. Being married, having given birth before, not being in government employment, and being older than 19 years were associated with late booking. Because the design was cross-sectional and the sample was non-random and from a single facility, these findings describe women attending this facility and show association, not causation.

## Declarations

### Ethics approval and consent to participate

The study was conducted in accordance with the principles of the Declaration of Helsinki. It was approved by the University of Zambia Biomedical Research Ethics Committee (UNZABREC; REF. No. 5212-2024) and the National Health Research Authority, and permission to collect data was obtained from the Mazabuka District Health Office. Women aged 18 years and above gave written informed consent. For girls aged 15 to 17 years, written parental or guardian consent was obtained and the girl gave written assent. Participants who could not write gave verbal consent with a thumbprint. Participation was voluntary.

### Consent for publication

Not applicable.

### Availability of data and material

The data set analyzed during the study is available from the author upon reasonable request.

### Competing interest

The author has no competing interests.

### Funding statement

The study was not funded.

### Author’s contribution

The author was responsible for the Conceptualization, Methodology, analyzing the data, interpreting the findings, and drafting the manuscript.

## Data Availability

The data set analyzed during the study is available from the author on reasonable request and can be accessed from the University of Zambia's main Library.

## Acknowledgement

The author wishes to send sincere appreciation and acknowledgments to the lecturers at the University of Zambia, School of Public Health, in particular Dr. Alice Ngoma Hazemba and Dr. Isaac Fwemba for the mentorship; Mazabuka District Health Office and Magoye Rural Health Center for the permission to collect data; NHRA for researcher certification; and UNZABREC for ethical approval granted.

## Author’s information

The research was part of a thesis by MCNM, who was pursuing a Master of Science in Public Health at the University of Zambia, School of Public Health.

## AI statement

During the preparation of this manuscript, Claude (Anthropic; Claude Sonnet 5.5, accessed October 2026) was used for language refinement, improvement in manuscript presentation, and drafting of the point-by-point reviewer response sheet and STROBE checklist. The author reviewed and verified all output and takes full responsibility for the content and conclusions of this manuscript.

## Notes

### Competing Interest Statement

The authors have declared no competing interest.

### Author Declarations

UNIVERSITY OF ZAMBIA BIOMEDICAL RESEARCH ETHICS AND NATIONAL HEALTH RESEARCH AUTHORITY

### Summary of Updates

work on errors pointed out by reviewers, format tables and work on the discussions and recommentions

## References

Banda, I. (2012) Factors associated with late antenatal care attendance in selected rural and urban communities of the Copperbelt province, Zambia. MPH dissertation. University of Zambia, Lusaka. Available at: https://dspace.unza.zm/items/43ed805a-e73c-4f29-8b32-b7cab8e506b6

Chewe, M.M. (2015) Factors associated with late antenatal care booking among pregnant women in Ndola, Zambia. MSc thesis. University of Zambia, Lusaka. Available at: https://dspace.unza.zm/handle/123456789/4285

Dorji, T., Das, M. and Van den Bergh, R. (2019) ‘“If we miss this chance, it’s futile later on” – late antenatal booking and its determinants in Bhutan: a mixed-methods study’, BMC Pregnancy and Childbirth, 19, article 158. Available at: 10.1186/s12884-019-2308-5

Ebonwu, J., Mumbauer, A., Uys, M., Wainberg, M.L. and Medina-Marino, A. (2018) ‘Determinants of late antenatal care presentation in rural and peri-urban communities in South Africa: a cross-sectional study’, PLOS ONE, 13(3), e0191903. Available at: 10.1371/journal.pone.0191903

Jinga, N., Mongwenyana, C., Moolla, A., Malete, G. and Onoya, D. (2019) ‘Reasons for late presentation for antenatal care, healthcare providers’ perspective’, BMC Health Services Research, 19, article 1016. Available at: 10.1186/s12913-019-4855-x

Mare, K.U., Andargie, G.G., Moloro, A.H. et al. (2025) ‘Late initiation of antenatal care visit amid implementation of new antenatal care model in Sub-Saharan African countries: a multilevel analysis of multination population survey data’, PLOS ONE, 20(1), e0316671. Available at: 10.1371/journal.pone.0316671

Ministry of Health (2012) HMIS instructions and procedures reference materials, Zambia. Lusaka: Ministry of Health.

Ministry of Health (2022) Annual progress report. Lusaka: Ministry of Health. Available at: https://www.moh.gov.zm/?wpfb_dl=231

Ndomba, A., Ntabaye, M., Semali, I., Kabalimu, T., Ndossi, G. and Mashalla, Y. (2023) ‘Prevalence of late antenatal care booking among pregnant women attending public health facilities of Kigamboni Municipality in Dar es Salaam region, Tanzania’, African Health Sciences, 23(2). Available at: 10.4314/ahs.v23i2.72

Olufemi Olayinka, T., Sebutu Bello, I., Oluwafemi Olajubu, T., Oloyede Oyegbade, O., Omobolanle Olajubu, A. and Tamunotonye Ezeoma, I. (2022) ‘Factors influencing the booking gestational age among antenatal clinic attendees at primary health centers in South West, Nigeria: a cross-sectional study’, SAGE Open Nursing, 8. Available at: 10.1177/23779608221139078

Pattinson, R.C. and Buchmann, E.J. (2017) Basic antenatal care plus handbook. 2nd edn. Pretoria: National Department of Health (SAMRC/University of Pretoria Maternal and Infant Health Care Strategies Research Unit). Available at: https://www.health.gov.za/wp-content/uploads/2023/05/BANC-Plus-Handbook-Feb-2017-with-NDoH-logo.pdf

Sinyange, N., Sitali, L., Jacobs, C., Musonda, P. and Michelo, C. (2016) ‘Factors associated with late antenatal care booking: population-based observations from the 2007 Zambia demographic and health survey’, Pan African Medical Journal, 25, article 109. Available at: 10.11604/pamj.2016.25.109.6873

Towongo, M.F., Ngome, E., Navaneetham, K. and Letamo, G. (2022) ‘Factors associated with women’s timing of first antenatal care visit during their last pregnancy: evidence from 2016 Uganda demographic health survey’, BMC Pregnancy and Childbirth, 22, article 829. Available at: 10.1186/s12884-022-05167-z

Warri, D. and George, A. (2020) ‘Perceptions of pregnant women of reasons for late initiation of antenatal care: a qualitative interview study’, BMC Pregnancy and Childbirth, 20, article 70. Available at: 10.1186/s12884-020-2746-0

World Health Organization (2016) WHO recommendations on antenatal care for a positive pregnancy experience. Geneva: World Health Organization. Available at: https://www.who.int/publications/i/item/9789241549912

World Health Organization (n.d.) Adolescent health. Available at: https://www.who.int/health-topics/adolescent-health

Zambia Statistics Agency, Ministry of Health (MoH) Zambia and ICF (2025) Zambia demographic and health survey 2024. Lusaka, Zambia, and Rockville, Maryland, USA: Zambia Statistics Agency, MoH, and ICF.

